# Humoral Immunity after mRNA Omicron JN.1 Vaccination

**DOI:** 10.1101/2024.09.04.24313057

**Authors:** Christine Happle, Markus Hoffmann, Amy Kempf, Inga Nehlmeier, Metodi V. Stankov, Noemi Calderon Hampel, Torsten Witte, Stefan Pöhlmann, Georg M. N. Behrens, Alexandra Dopfer-Jablonka

**Affiliations:** Department of Rheumatology and Immunology, Hannover Medical School, Hannover, Germany; Department of Pediatric Pulmonology, Allergology and Neonatology, Hannover Medical School, Hannover, Germany; Infection Biology Unit, German Primate Center – Leibniz Institute for Primate Research, Göttingen, Germany; Faculty of Biology and Psychology, Georg-August-University Göttingen, Göttingen, Germany; Cluster of Excellence RESIST (EXC 2155), Hannover Medical School, 30625 Hannover, Germany; Center for Individualized Infection Medicine (CiiM), Hannover, Germany

## Abstract

In late June 2024, the European Medicines Agency (EMA) recommended market authorization for a monovalent COVID-19 mRNA-vaccine based on JN.1 spike. We assessed immune responses in n=42 health-care workers (median age 47 years, interquartile range, IQR 19·5 years, 48% male), who in August 2024 were vaccinated with 30 μg of the updated mRNA omicron JN.1 vaccine (bretovameran, BioNTech/Pfizer, Mainz, Germany). Humoral immune responses were analyzed directly prior to and 13 days after vaccination.

The omicron JN.1 vaccination resulted in a significant 1·2-fold increase of anti-S IgG and 1·2-fold increase of omicron anti-S IgG (p<0·0001). To assess plasma neutralisation capacity, we employed a pseudovirus particle (pp) neutralisation assay including S proteins of seven SARS-CoV-2 lineages. Baseline response rates were 100% for XBB.1.5_pp_, 90% for JN.1_pp_ and KP.2_pp_, 82% for KP.2.3_pp_, 92% for KP.3_pp_, and 72% for LB.1_pp_. Before JN.1 vaccination, particles bearing KP sublineage S proteins were slightly less efficiently neutralised compared with JN.1_pp_ (median change, 1·2-fold to 2·6-fold), while LB.1_pp_ neutralisation was 3-fold reduced, indicating antibody evasion. After vaccination, the response rates increased significantly for all pseudoviruses except XBB.1.5_pp_ and KP.3_pp_. Thus, we observed a significant increase in neutralisation of JN.1_pp_, KP.2_pp_, KP.2.3_pp_, and LB.1_pp_, showing a median change of 2.2-fold, 3.8-fold, 3.3-fold, and 4.9-fold, respectively.

In summary, bretovameran increased anti-S IgG and strengthened neutralising responses against circulating SARS-CoV-2-variants, except for KP.3. We wish to point out that our study population exhibited high pre-vaccination omicron-related hybrid immunity and may not be representative of other populations. Our data supports the notion that the new mRNA vaccine against omicron JN.1 most likely increases protection against hospitalization and post-COVID sequelae caused by most current variants.

---

The constant emergence of SARS-CoV-2 variants and sublineages that evaded control by neutralising antibodies induced upon infection and/or vaccination required the use of adapted vaccines. A vaccine adapted to the XBB1.5 variant became available in autumn of 2023 and initially provided robust protection against hospitalization due to infection [1,2]. The omicron JN.1 lineage, which became dominant in 2024, is currently being replaced by JN.1 sublineages including KP.2 and KP.3. However, protection installed by the XBB.1.5 adapted vaccine is probably inefficient at present, due to waning antibody titers. Therefore, in late June 2024, the European Medicines Agency (EMA) recommended market authorization for a monovalent COVID-19 mRNA-vaccine based on JN.1 spike [4]. However, immune response data in humans or real-world evidence on vaccine-induced protection is pending. To generate such data, we assessed immune responses in n=42 health-care workers (median age 47 years, interquartile range, IQR 19·5 years, 48% male), who in August 2024 were vaccinated with 30 μg of the updated mRNA omicron JN.1 vaccine (bretovameran, BioNTech/Pfizer, Mainz, Germany). Humoral immune responses were analyzed directly prior to and 13 days after vaccination. The median number of previous COVID-19 vaccinations was 4·5 (IQR 1), 87·8% of vaccinees reported at least one previous SARS-CoV-2 infection, and 97·6 % of the participants had been exposed to omicron antigen (appendix p.2). We independently assessed n=14 study participants with breakthrough infections caused by contemporary SARS-CoV-2 variants circulating in July and August 2024 in Germany.

First, we determined SARS-CoV-2 anti-spike (anti-S) IgG antibodies prior to and after JN.1 vaccination. Before JN.1 immunisation, participants showed a median of 2217 antibody-binding units per mL (IQR 2709) of anti-S IgG antibodies and a median of 338 relative units per mL (IQR 348·5) of omicron IgG antibodies (figure A), which was about twice as high as compared to immune responses prior to the omicron-directed vaccination against XBB.1.5 we had previously observed [5]. The omicron JN.1 vaccination resulted in a significant 1·2-fold increase of anti-S IgG and 1·2-fold increase of omicron anti-S IgG (p<0·0001, figure A). The absolute increase in both anti-S IgG types after JN.1 vaccination was comparable to that measured after vaccination with the XBB.1.5 adapted vaccine [5].

To assess plasma neutralisation capacity, we employed a pseudovirus particle (pp) neutralisation assay (ie, pseudovirus neutralisation test), including S proteins of seven SARS-CoV-2 lineages (figure B, appendix pp 8). Baseline response rates were 100% for XBB.1.5_pp_, 90% for JN.1_pp_ and KP.2_pp_, 82% for KP.2.3_pp_, 92% for KP.3_pp_, and 72% for LB.1_pp_ (figure B). Before JN.1 vaccination, particles bearing KP sublineage S proteins were slightly less efficiently neutralised compared with JN.1_pp_ (median change, 1·2-fold to 2·6-fold), while LB.1_pp_ neutralisation was 3-fold reduced, indicating antibody evasion; figure C). After vaccination, the response rates increased significantly for all pseudoviruses except XBB.1.5_pp_ and KP.3_pp_ (figure C, appendix p10). Thus, we observed a significant increase in neutralisation of JN.1_pp_, KP.2_pp_, KP.2.3_pp_, and LB.1_pp_, showing a median change of 2.2-fold, 3.8-fold, 3.3-fold, and 4.9-fold, respectively. Postvaccination neutralisation titers were within comparable ranges to individuals with recent breakthrough infections caused by contemporary variants (appendix p11).

In summary, bretovameran, an mRNA vaccine adapted to the spike protein of the omicron JN.1 variant, increased anti-S IgG in all vaccinated persons at 13 days post vaccination and strengthened neutralising responses against circulating SARS-CoV-2-variants, except for KP.3. Our data are in concordance with preclinical findings with fourfold vaccinated mice [6] and expand our knowledge on JN.1-associated immunity. We wish to point out that our study population exhibited high pre-vaccination omicron-related hybrid immunity, which could impact the magnitude and quality of humoral immunity induced by the JN.1-adpated vaccine and may not be representative of other populations. Despite this and some limitations listed in detail in the appendix (p 5), our data supports the notion that the new mRNA vaccine against omicron JN.1 most likely increases protection against hospitalization and post-COVID sequelae caused by most current variants.

1. Hansen CH, Moustsen-Helms IR, Rasmussen M, Soborg B, Ullum H, Valentiner-Branth P: Short-term effectiveness of the XBB.1.5 updated COVID-19 vaccine against hospitalisation in Denmark: a national cohort study. Lancet Infect Dis 2024, 24(2):e73-e74.
2. Ma KC, Surie D, Lauring AS, Martin ET, Leis AM, Papalambros L, Gaglani M, Columbus C, Gottlieb RL, Ghamande S, Peltan ID, Brown SM, Ginde AA, Mohr NM, Gibbs KW, Hager DN, Saeed S, Prekker ME, Gong MN, Mohamed A, Johnson NJ, Srinivasan V, Steingrub JS, Khan A, Hough CL, Duggal A, Wilson JG, Qadir N, Chang SY, Mallow C, Kwon JH, Parikh B, Exline MC, Vaughn IA, Ramesh M, Safdar B, Mosier J, Harris ES, Shapiro NI, Felzer J, Zhu Y, Grijalva CG, Halasa N, Chappell JD, Womack KN, Rhoads JP, Baughman A, Swan SA, Johnson CA, Rice TW, Casey JD, Blair PW, Han JH, Ellington S, Lewis NM, Thornburg N, Paden CR, Atherton LJ, Self WH, Dawood FS, DeCuir J. Effectiveness of Updated 2023-2024 (Monovalent XBB.1.5) COVID-19 Vaccination Against SARS-CoV-2 Omicron XBB and BA.2.86/JN.1 Lineage Hospitalization and a Comparison of Clinical Severity-IVY Network, 26 Hospitals, October 18, 2023-March 9, 2024. Clin Infect Dis. 2024 Aug 6:ciae405. doi: 10.1093/cid/ciae405. Epub ahead of print. PMID: 39107255.
3. Zhang L, Dopfer-Jablonka A, Cossmann A, Stankov MV, Graichen L, Moldenhauer AS, Fichter C, Aggarwal A, Turville SG, Behrens GMN et al: Rapid spread of the SARS-CoV-2 JN.1 lineage is associated with increased neutralization evasion. iScience 2024, 27(6):109904.
4. European Medical Association Emergency Task Force: EMA confirms its recommendation to update the antigenic composition of authorised COVID-19 vaccines for 2024-2025. 2024 (https://www.ema.europa.eu/en/documents/other/ema-confirms-its-recommendation-update-antigenic-composition-authorised-covid-19-vaccines-2024-2025_en.pdf), *last accessed 20240904*.
5. Stankov MV, Hoffmann M, Gutierrez Jauregui R, Cossmann A, Morillas Ramos G, Graalmann T, Winter EJ, Friedrichsen M, Ravens I, Ilievska T, Ristenpart J, Schimrock A, Willenzon S, Ahrenstorf G, Witte T, Förster R, Kempf A, Pöhlmann S, Hammerschmidt SI, Dopfer-Jablonka A, Behrens GMN. Humoral and cellular immune responses following BNT162b2 XBB.1.5 vaccination. Lancet Infect Dis. 2024 Jan;24(1):e1-e3.
6. Biontech/Pfizer_Vaccines_and_Related_Biological_Products_Advisory¬_Committee: Vaccines and Related Biological Products Advisory Committee June 5, 2024 Meeting Presentation-Pfizer/BioNTech Clinical and Preclinical Supportive Data 2024-2025 COVID19 Vaccine Formula. https://wwwfdagov/media/179144/download 2024, *last accessed 20240904*.

**Figure:**
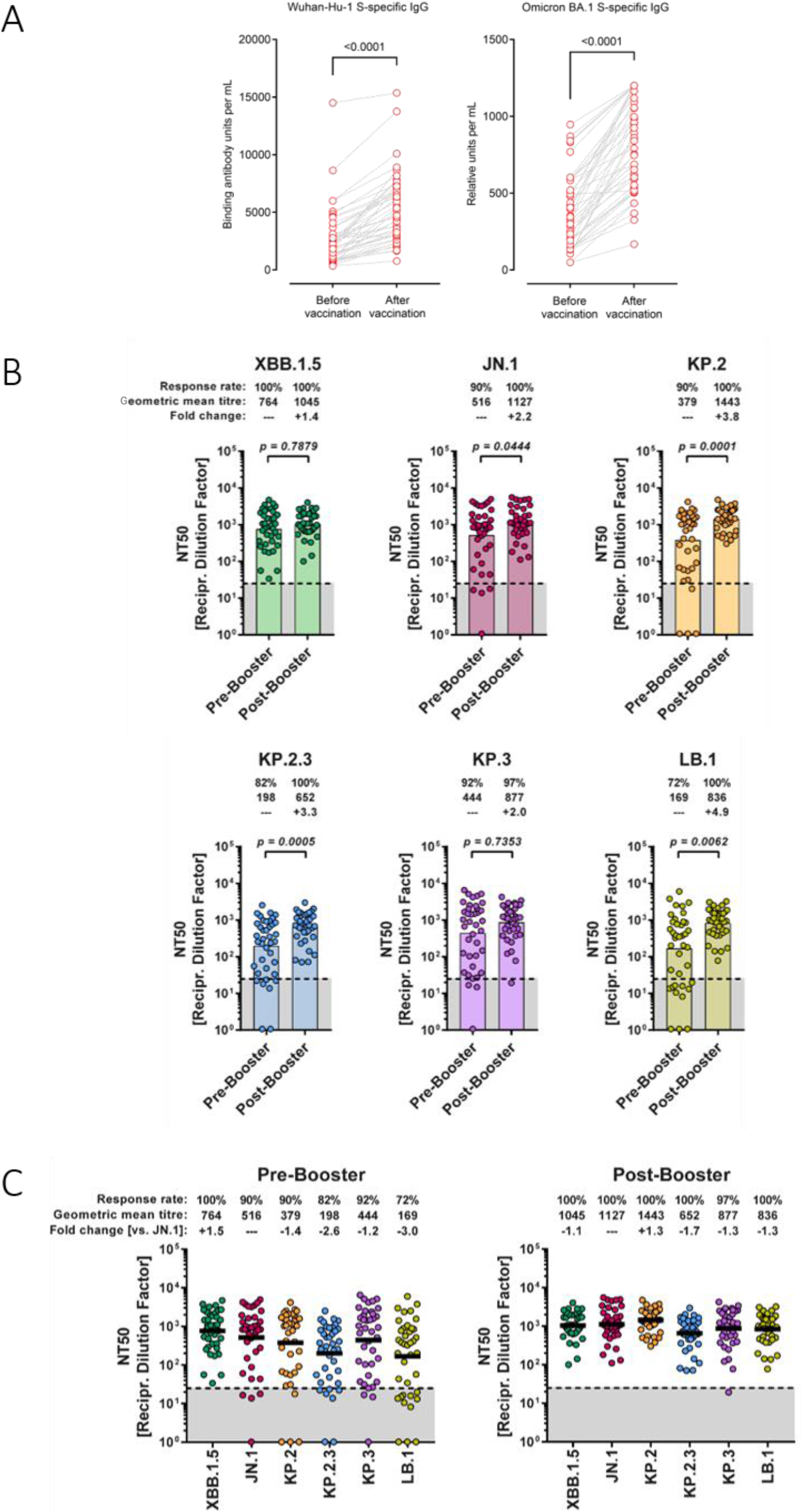
Humoral immune responses following mRNA omicron JN.1 vaccination. (A) Concentrations of Wuhan-Hu-1 S-specific IgG and omicron S-specific IgG in plasma obtained before or after vaccination with the mRNA omicron JN.1 vaccine. (B) Neutralisation of pseudovirus particles bearing the indicated S proteins by donor-matched plasma (n=39) taken before or after vaccination with the mRNA omicron JN.1 vaccine. Data represent GMT (colored columns) from a single experiment, performed with four technical replicates. The lowest plasma dilution tested (dashed lines) and the threshold (lower limit of detection; grey shaded areas) are indicated. Information on response rates and median fold change in neutralisation after vaccination are indicated above the graphs. Of note, for graphical reasons, plasma samples yielding an NT50 value below 6·25 (limit of detection) were manually set at bottom of the axis. (C) The data presented in panel C were regrouped to compare differences in SARS-CoV-2 lineage-specific neutralisation before and after vaccination. Information on GMT (also indicated by horizontal lines) response rates, and median fold change in neutralisation compared with JN.1 pseudovirus particles are indicated above the graphs. Individual neutralisation data are available in the appendix (pp 14–15). NA=not applicable. S=spike. GMT=geometric mean titres.

## Data Availability

All requests for raw and analysed data that underly the results reported in this article will be reviewed within four weeks by the CoCo Study Team, Hannover Medical School to determine whether the request is subject to confidentiality and data protection obligations. Data that can be shared will be released via a material transfer agreement.

## Appendix

**Table S1.**
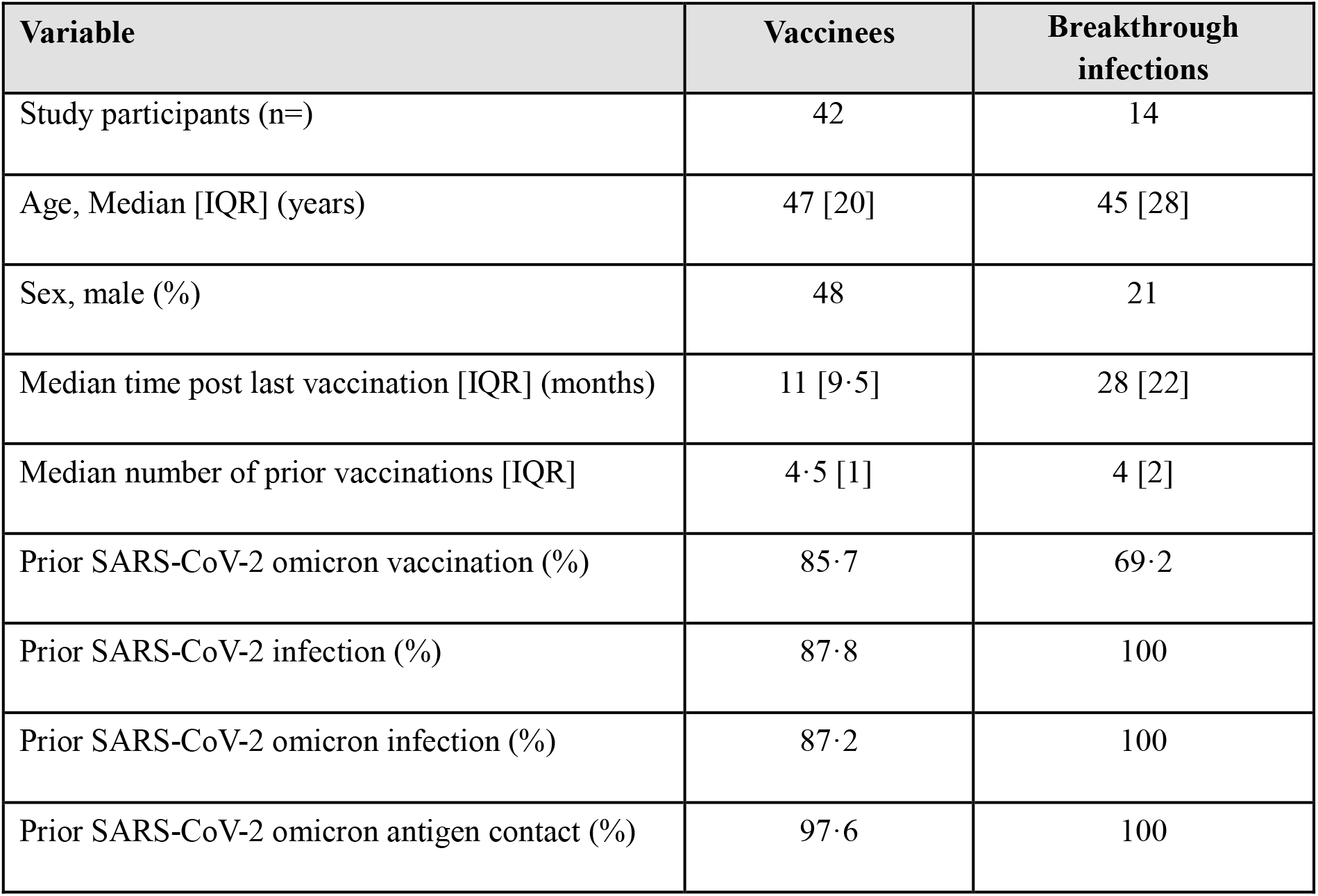
Demographics, and infection and vaccination history.

| **Variable** | **Vaccinees** | **Breakthrough infections** |
| --- | --- | --- |
| Study participants (n=) | 42 | 14 |
| Age, Median [IQR] (years) | 47 [20] | 45 [28] |
| Sex, male (%) | 48 | 21 |
| Median time post last vaccination [IQR] (months) | 11 [9·5] | 28 [22] |
| Median number of prior vaccinations [IQR] | 4·5 [1] | 4 [2] |
| Prior SARS-CoV-2 omicron vaccination (%) | 85·7 | 69·2 |
| Prior SARS-CoV-2 infection (%) | 87·8 | 100 |
| Prior SARS-CoV-2 omicron infection (%) | 87·2 | 100 |
| Prior SARS-CoV-2 omicron antigen contact (%) | 97·6 | 100 |

## Methods

### Participants

For this study, we recruited n=46 individuals from the COVID-19 Contact (CoCo) Study (German Clinical Trial Registry, DRKS00021152) that were vaccinated with Comirnaty^®^ omicron JN.1/bretovameran and analysed n=42 individuals for which 13-day follow-up data was available and no SARS-CoV-2 infection between vaccination and day 13 was reported. The CoCo study is an ongoing, prospective, observational study monitoring anti-SARS-CoV-2 immunoglobulin G and immune responses in healthcare professionals at Hannover Medical School [1]. In August 2024, CoCo study participants that decided to receive a COVID-19 vaccination with bretovameran as part of the German vaccination campaign, were asked to donate blood before and after vaccination. In addition, we included n=14 participants with recent COVID-19 in July or August 2024 confirmed by quick test for comparison (all anti-nucleocapsid protein (NCP) IgG positive). According to the German wastewater-based surveillance on SARS-CoV-2, KP.3.1.1, LB.1, KP.3, KP.2 and JN.1 were among the most detected variants [2].

We estimated that a sample size of n=42 would be sufficient to detect a clinically meaningful difference within the group, assuming that S protein-specific IgG levels double from pre-vaccination (mean 822 (SD 747) RU/mL, after: mean 1644 RU/mL (SD 1,494). This estimation was based on anti-S IgG measurements in a convenience sample of 24 persons from the CoCo cohort in August 2023, which is our best estimate of pre-vaccination levels, correlation between groups 0·5. This estimate is based on a one-tailed paired t-test of differences between means, with 95% power and 1% level of significance. From our previous experiences, we estimated a loss-to follow-up rate of 10%. Therefore, a sample size of 46 vaccinated persons has been aimed at. The power calculation was performed using G*Power, Version 3.1.9.6.

Participants that were vaccinated with bretovameran were invited to donate blood before and 13 days after vaccination at which time robust antibody responses are detectable [3]. The CoCo Study cohort comprises a general population of health-care professionals, without specific pre-existing conditions. Twenty-seven percent of the bretovameran vaccinees reported underlying conditions (such as e.g. asthma), and three participants reported treatment with either methotrexate, ixekizumab plus sulfasalazin, or upadacitinib. No individual developed positive anti-NCP IgG after vaccination. Demographics (sex and age), infection, and vaccination history, respectively, are depicted in Table S1.

### Serology

Serology was performed essentially as described before [3]. We separated plasma from lithium heparin blood (S-Monovette, Sarstedt) and stored it at 4 °C for immediate use or at -20 °C until use. We measured SARS-CoV-2 IgG by quantitative ELISA (anti-SARS-CoV-2 S1 Spike protein domain/receptor binding domain IgG SARS-CoV-2-QuantiVac, EI 2606-9601-10G, and S1 Spike protein domain/receptor binding domain IgG SARS-CoV-2 of omicron, EI 2606-9601-30 G, both EUROIMMUN, Lübeck, Germany) according to manufacturer’s instructions (dilution up to 1:4,000). We used anti-S1 concentrations expressed as RU/mL as assessed from a calibration curve with values above 11 RU/mL defined as positive [1] and provide results in binding antibody units (BAU/mL). We performed anti-NCP IgG measurements according to the manufacturer’s instructions (EUROIMMUN, Lübeck, Germany) and used an AESKU.READER (AESKU.GROUP, Wendelsheim, Germany) and the Gen5 2.01 Software for analysis.

### Production of pseudovirus particles and pseudovirus neutralisation test (pVNT)

pVNTs were conducted at the Infection Biology Unit of the German Primate Centre in Göttingen according to a previously published protocol^2^ with minor modifications. The following S protein expression plasmids were used: pCG1_SARS-2-SΔ18 XBB.1.5 (EPI_ISL_16239158; codon-optimised, deletion of last 18 aa residues at the C-terminus) [4], pCG1_SARS-2-SΔ18 JN.1 (EPI_ISL_18530042; codon-optimised, deletion of the last 18 aa residues at the C-terminus) [5], pCG1_SARS-2-SΔ18 KP.2 (EPI_ISL_19197864; codon-optimised, deletion of the last 18 aa residues at the C-terminus), pCG1_SARS-2-SΔ18 KP.2.3 (EPI_ISL_19197559; codon-optimised, deletion of the last 18 aa residues at the C-terminus), pCG1_SARS-2-SΔ18 KP.3 (EPI_ISL_19203001; codon-optimised, deletion of last 18 aa residues at the C-terminus), pCG1_SARS-2-SΔ18 LB.1 (EPI_ISL_19067004; codon-optimised, deletion of the last 18 aa residues at the C-terminus). Expression plasmids for KP.2, KP.2.3, KP.3, and LB.1 S proteins were generated by introduction of the respective mutations into plasmid pCG1_SARS-2-SΔ18 JN.1 through overlap-extension PCR with overlapping primers harbouring the desired mutations.

For the production of pseudovirus particles harbouring S proteins of the SARS-CoV-2 lineages under study, 293T cells expressing the respective S protein following transfection were inoculated with VSV*DG-FLuc, ^9^ a replication-deficient VSV vector that lacks the genetic information for the VSV glycoprotein and instead encodes for an enhanced green fluorescent protein and a firefly luciferase (FLuc) (kindly provided by Gert Zimmer, Institute of Virology and Immunology, Mittelhäusern, Switzerland). After an incubation period of 1 h at 37 °C, cells were washed with PBS and further incubated with medium containing anti-VSV-G antibody (culture supernatant from I1-hybridoma cells; ATCC no. CRL-2700) to neutralise residual input virus. At 16-18 h post inoculation, pseudovirus particles were harvested. For this, the culture medium was centrifuged (4,000 x g, 10 min) and clarified supernatants were aliquoted and stored at -80 °C until further use.

pVNTs were performed using Vero76 cells (kindly provided by Andrea Maisner, Institute for Virology, Phillips University Marburg) that were seeded in 96-well plates. Before analysis, all plasma samples were heat-inactivated (56 °C, 30 min) and serially diluted in culture medium. Next, equal volumes of the serially diluted plasma samples (final dilution range 1:25 to 1:6,400) and pseudovirus particles were mixed and incubated for 30 min at 37 °C, before the mixtures were inoculated onto confluent Vero76 monolayers. Of note, pseudovirus particles incubated with medium alone instead of plasma sample served as reference. Pseudovirus infection was analysed at 16-18 h post inoculation by measuring FLuc activity in cell lysates. For this, the cell culture supernatant was aspirated before the cells were lysed with PBS containing 0·5 % Tergitol (Carl Roth; 30 min at room temperature). Subsequently, cell lysates were transferred into white 96-well plates and mixed with FLuc substrate (Beetle-Juice, PJK), before luminescence was recorded using a Hidex Sense Microplate Reader Software (version 0.5.41.0).

Neutralisation efficiency was determined based on the relative inhibition of pseudovirus entry, with signals obtained from pseudovirus particles incubated in the absence of plasma serving as reference (= 0% inhibition). In addition, we used a non-linear regression model to calculate the neutralising titre 50 (NT50), indicating the plasma dilution required for half-maximal inhibition of pseudovirus infection. Of note, plasma samples that yielded NT50 values below 25 were considered as non-responders. Further, plasma samples that yielded NT50 values below 6·25 (limit of detection, LOD) were assigned an NT50 value of 3·125 (0.5 of LOD). Two samples that yielded NT50 values above 6400 before vaccination were excluded from the analysis, as they were out of the quantifiable range of this assay and one sample was missing for pairwise pVNT analysis.

### Statistics

Statistical analysis was conducted using GraphPad Prism 8.4 or 9.0 (GraphPad Software, USA) and SPSS 20.0.0 (IBM SPSS Statistics, USA). Outliers were included in the analysis, and missing values were excluded pairwise. Mean (SD) was used for normally distributed data, while median (IQR) was used for non-normally distributed data. In non-normally distributed data, Wilcoxon-Mann-Whitney-tests were used. Neutralisation titres were transformed to geometric mean titres.

## Limitations of the study

Our study has some limitations. Although our previous analysis [3] confirmed that anti-S IgG and neutralising antibodies plateaued at day eight to ten post omicron XBB.1.5 vaccination, which is in keeping with antibody kinetics described after the second BNT162b2 vaccination [6] or after other COVID-19 vaccinations [7], our data are preliminary and humoral and cellular immunity could further strengthen over time. Furthermore, this data can only provide first insights into the initial immune response to the updated JN.1 vaccine. Longitudinal data will be necessary to assess immune trends and durability. In addition, most of our vaccinees had previous SARS-CoV-2 Omicron infections and/or multiple vaccinations, likely contributing to the considerable anti-S IgG response present already before mRNA omicron JN.1 vaccination. Finally, neutralisation of SARS-CoV-2 lineages was assessed by pVNT, which has been shown to serve as an adequate surrogate model for this purpose [8]. Nevertheless, our data formally await confirmation with clinical isolates and eventually validation in studies with clinical endpoints.

**Acknowledgements**

## Contributions

Study design: G.M.N.B., A.D.-J.

Data collection: C.H., N.C.H, A.K., I.N.

Data curation: C.H., N.C.H

Data analysis: G.M.N.B, A.D.J., C.H., M.H., M.V.S., T.W.

Data interpretation: M.H., S.P., G.M.N.B., A.D.-J.

Writing: C.H., G.M.N.B. with comments from all authors.

G.M.N.B and A.D.-J. have directly accessed and verified the underlying data reported in the manuscript.

## Ethics committee approval

The CoCo Study (German Clinical Trial Register DRKS00021152) and the analysis conducted for this article were approved by the Internal Review Board of Hannover Medical School (institutional review board no. 8973_BO-K_2020, last amendment Aug 2024). All study participants gave written informed consent and received no compensation.

## Acknowledgements

G.M.N.B. and A.D.-J. acknowledge funding (Niedersächsisches Ministerium für Wissenschaft und Kultur; 14-76103-184, COFONI Network, project 4LZF23), G.M.N.B. acknowledge funding by the European Regional Development Fund ZW7-85151373, and A.D.-J. acknowledge funding by European Social Fund (ZAM5-87006761). T.W. acknowledges funded by the DFG under Germany’s Excellence Strategy EXC 2155 “RESIST” (Project 390874280). S.P. acknowledges funding by the EU project UNDINE (grant agreement number 101057100), the COVID-19-Research Network Lower Saxony (COFONI) through funding from the Ministry of Science and Culture of Lower Saxony in Germany (14-76103-184, projects 7FF22, 6FF22, 10FF22) and the German Research Foundation (Deutsche Forschungsgemeinschaft, DFG; PO 716/11-1). The funding sources had no role in the design and execution of the study, the writing of the manuscript and the decision to submit the manuscript for publication. The authors did not receive payment by a pharmaceutical company or other agency to write the publication. The authors were not precluded from accessing data in the study, and they accept responsibility to submit for publication.

We thank the CoCo Study participants for their support and the entire CoCo study team for help. We would like to thank Janine Topal, Kerstin Sträche, Birgit Heinisch, Andrea Stölting, Simon F. Ritter, Louis Kuhnke, Eva Stöppelmann, Melanie Ignacio, and Marion Hitzigrath for technical and logistical support.

---

**Fig. S1.**
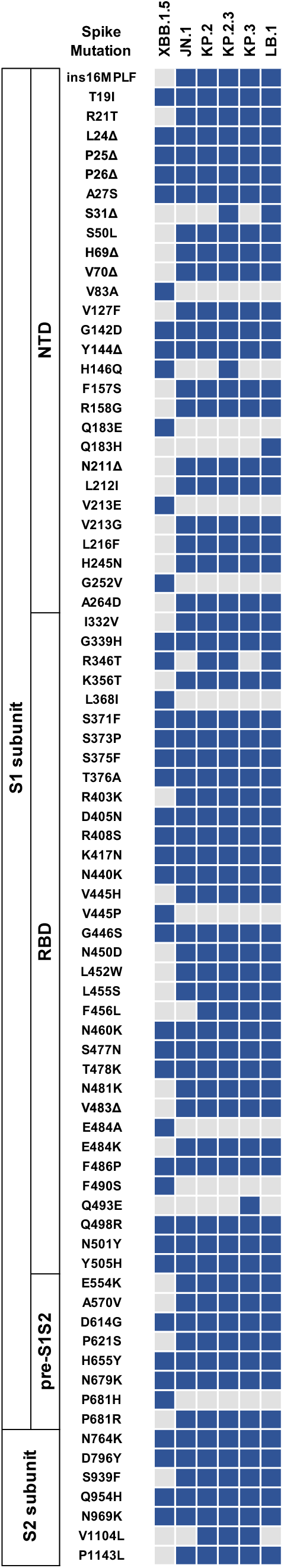
Overview of SARS-CoV-2 lineage-specific spike protein mutations.

**Fig. S2.**
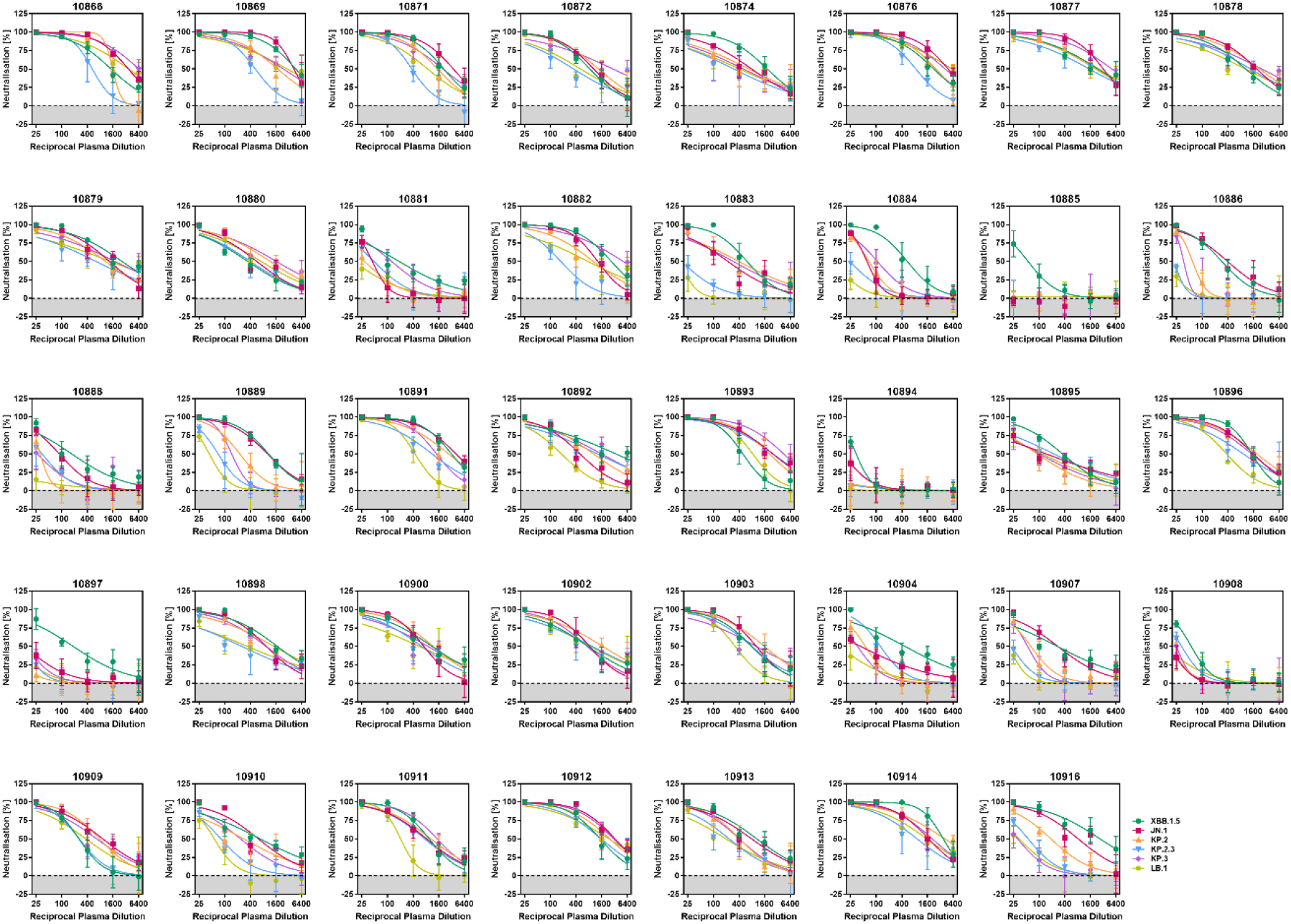
Individual neutralisation data for pre-vaccination plasma.

**Fig. S3.**
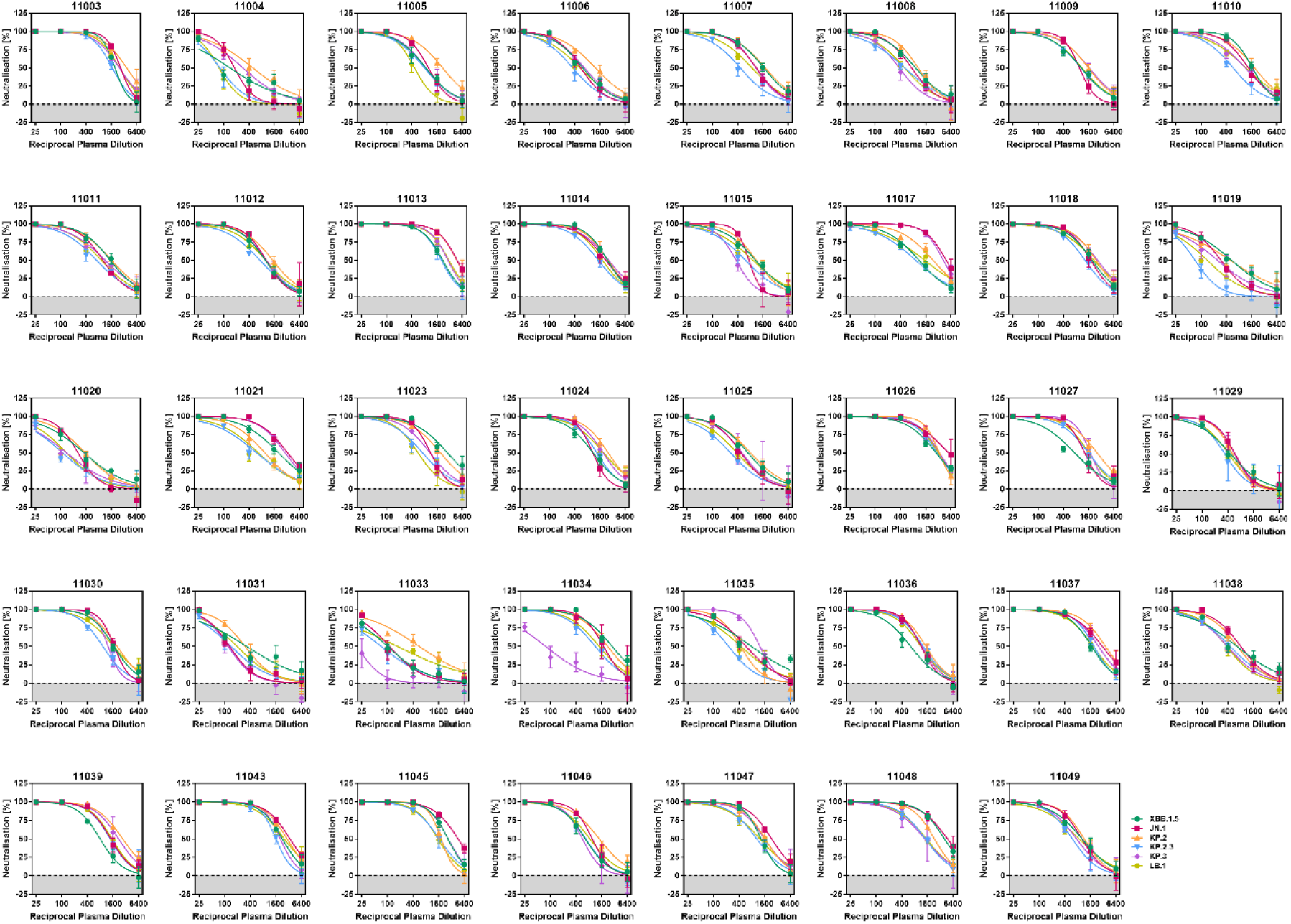
Individual neutralisation data for post-vaccination plasma.

**Fig. S4.**
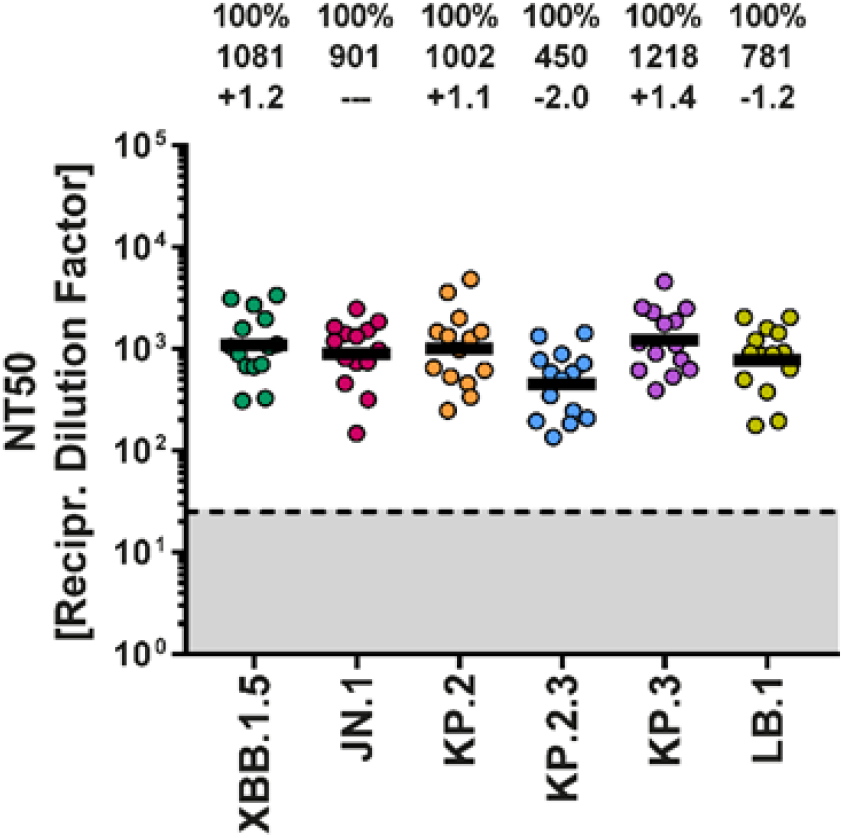
Neutralisation data for individuals with SARS-CoV-2 breakthrough infections in July and August 2024.

